# Rapid feedback on my Emergency Department Hemorrhagic Stroke care? It Improves Telestroke and In-person treatment times

**DOI:** 10.1101/2025.08.15.25333814

**Authors:** Nick M. Murray, Paul D. Johnson, Gabriel V. Fontaine, Trina Johnson, Jami Hassler, Heidi Hafen, Michelle Aucoin, Lesley Miller, Anne M. Anderson, Chamonix Johnston, Marilyn McKasson, Kyle Hobbs, H. Adrian Püttgen

**Author notes:** <u>Corresponding author contact information</u> Nick Murray MD, 5121 Cottonwood Street, Murray, UT, 84107. The authors of this original research confirm that the manuscript complies with all instructions to authors. All authorship requirements have been met, and the final manuscript was approved by all authors. This original research has not been published elsewhere and is not under consideration by another journal. Reporting checklist STROBE was used and uploaded to the submission portal. COI forms are complete for all authors and there are no relevant disclosures. Author contributions: NM and PJ developed study conception and design, data acquisition, data analysis, and manuscript writing. GVF, TJ, JH, HH, MA, LM, AMA, CJ, MM, KH, and HAP directed the study intervention in real-time, made key decisions in data analysis, and manuscript writing. This research adheres to ethical guidelines and was reviewed by the institutional review board (IRB), and a waiver of consent was approved. There were no funding sources. Artificial intelligence was not used for any portion of the manuscript.

## Abstract

**Background:** Rapid time to treatment of intracerebral hemorrhage (ICH) is important yet not consistently achieved in some emergency departments (EDs). The concept of CODE-ICH, similar to Code Ischemic Stroke, is now well-described, yet mechanisms to change provider and system behavior to improve treatment times is lacking.

**Objective:** We aim to evaluate ICH treatment time metrics before and after a standardized ICH provider feedback intervention, individualized for each patient with a spontaneous acute ICH in the ED.

**Methods:** A multicenter retrospective cohort of consecutive patients ≥ 18 years old with acute ICH were identified from March 1, 2022 to January 1, 2025 within a network of an integrated not-for profit healthcare system in the U.S. A rapid feedback intervention for all providers involved in the patient case was developed to contain specific treatment times compared to goals. Patients were grouped into pre- and post-intervention cohorts. The primary endpoint was CT scout time to antihypertensive agent and anticoagulation reversal agent administration. Secondary outcomes were: ICH orderset utilization, in-person daytime versus telehealth nighttime coverage, length of stay, and discharge disposition.

**Results:** A total of 226 patients met inclusion criteria, 108 pre- and 118 post-intervention, with similar age (median: 68 vs. 69 years) and 54% were female. Pre- to post-intervention median NIHSS was the same (10; p=0.25), as were median ICH scores (pre: 2.0, interquartile range, IQR, 0-3 vs. post: 1.0, IQR 1-3; p=0.90). Median post-intervention CT to antihypertensive treatment was faster (pre: 21, IQR 23-52 min vs. Post: 14, IQR 7-26 min; p=0.0012), as well as median CT to anticoagulation reversal agent administration (pre: 40, IQR 30-64 min vs. Post: 29, IQR 18-40 min; p=0.03). The intervention was associated with increased orderset usage (54% to 96%; p=0.0001).

**Conclusions:** A standardized ICH feedback intervention improved treatment times for blood pressure and anticoagulation reversal following detection of ICH on CT head imaging in the ED.

## INTRODUCTION

Rapid treatment is recommended for all patients with acute stroke symptoms that present to the emergency department (ED), including spontaneous intracerebral hemorrhage (ICH).(1)(2)(3) The time to treatment paradigm for ICH is exemplified by the bundled care concept of the INTERACT-3 trial, CODE-ICH analyses, and includes rapid blood pressure control, anticoagulation reversal, as well as glucose and fever control.(4)(5) However, implementation of these bundled interventions in a time sensitive manner is inconsistent.(6) Specific strategies or behavioral interventions to improve protocolized time-sensitive ICH care decisions are sparse.

Defined care pathways with a focus on reducing treatment time are well defined in ischemic stroke, where process improvement interventions have steadily reduced door to needle (DTN) time for intravenous (IV) thrombolytics. Interventions include: stroke prenotification, rapid patient registration, moving directly to CT scanning, giving IV thrombolytics in the CT scanner, and rapid feedback.(7)(8) Many of these practice paradigms have benefited the care of ischemic strokes, yet since ICH strokes do not follow the same treatment pathway after the CT scan, a more nuanced disease specific approach is needed. For example, inclusion of pharmacist teams specifically has shown improvement for ischemic stroke,(9)(10) and pharmacy directed care is a core component of basic time-sensitive ICH hypertension treatment and anticoagulation reversal.

Our multi-center hospital system has an established bundled care pathway for patients presenting to the emergency department with ICH. The primary aim is to evaluate if rapid feedback to a multidisciplinary team on CODE ICH performance and patient outcome improves time sensitive treatment metrics in ICH care.

## METHODS

### Study Design

We performed an IRB approved retrospective review of prospectively collected multi-institutional databases from a large, integrated, not-for-profit health system. Four hospitals were included: one comprehensive and three primary or thrombectomy capable stroke centers. Each hospital had standardized hemorrhagic stroke treatment protocols as well as an orderset in the electronic medical record (EMR), however the timing for introduction of the feedback intervention was variable between the sites, though all remained within a 6-month time frame. The patients at each site were grouped into pre- and post-intervention groups for each hospital.

### ICH Orderset and Post-ICH Feedback Intervention

At each of the four hospital sites, a standard ED ICH orderset was implemented. The orderset contained ICH-specific vital sign monitoring parameters, nursing neurological checks, labs, patient blood pressure and care goals, NPO status, swallow screen, antihypertensive medications, and anticoagulation reversal medications, with links to real-time guidance protocols.

The post-ICH rapid feedback intervention was then implemented at each site within a 6-month period. Patients pre- and post-intervention go live date at each site were combined into the appropriate, respective groups. The intervention specifically was based on a similar feedback form used for patients who received IV-thrombolytics and/or endovascular therapy in the setting of ischemic stroke, yet modified to show ICH specific time goals. The form contained the patient demographics and clinical course, and displayed patient specific treatment time metrics, which were coupled with recommended treatment goals. If the patient did not meet goals, this triggered a review process led by the stroke coordinator, and if appropriate, an individual goal-specific root cause analysis. This feedback form, in format of a PowerPoint slide JPEG was sent to all involved caregivers, including: emergency medical field staff, ED providers, nurses, CT technicians, pharmacists, neurologists, and the admitting team providers within 1-3 days of patient care (Figure 1).

**Figure 1.**
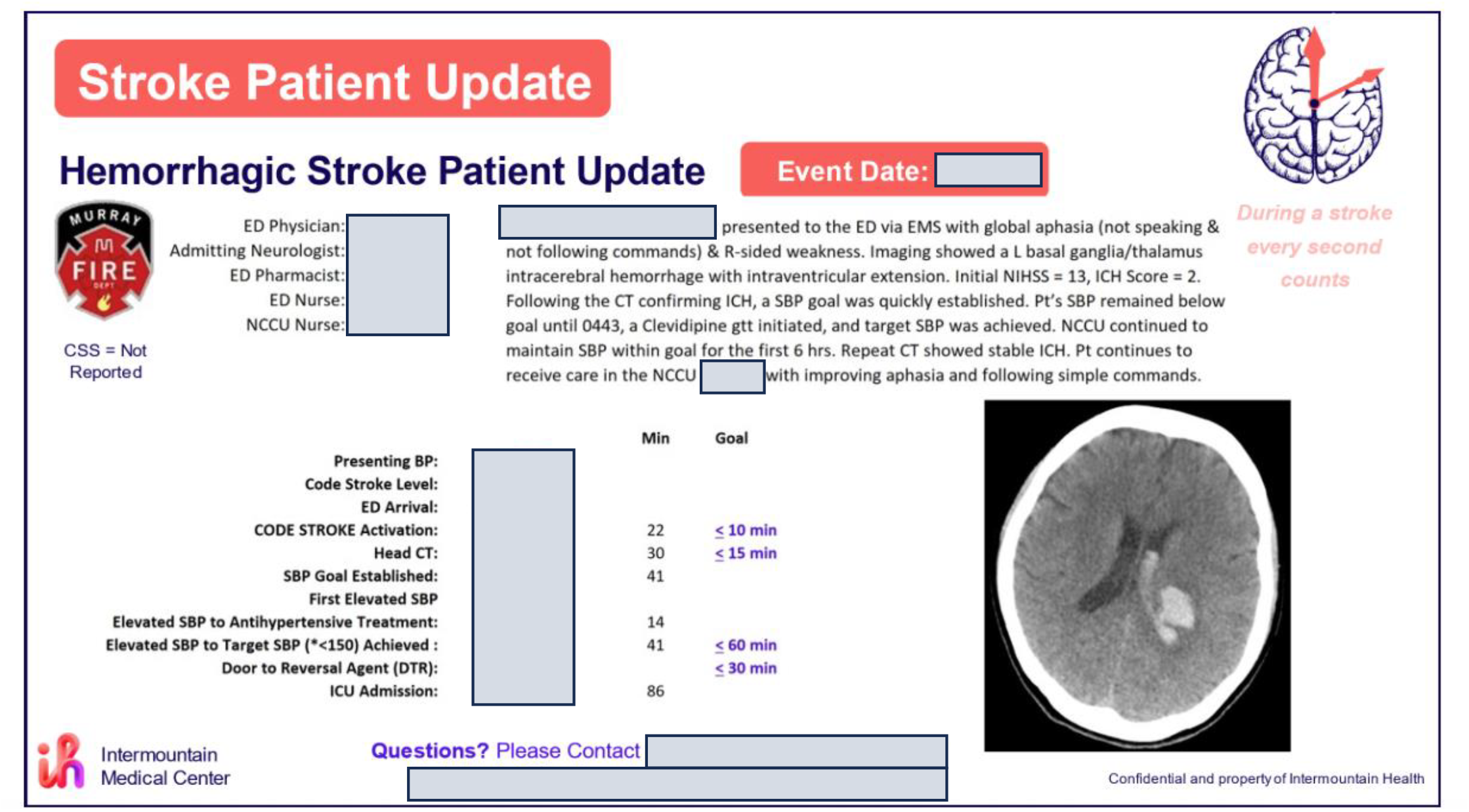
Feedback intervention form for ICH patients that was distributed to all providers and caregivers involved in the patient’s ED course.

### Patient Selection

Patients ≥ 18 years old who presented to the emergency ED as a stroke activation between March 1, 2022 to January 1, 2025 with non-contrast head CT imaging with primary finding of spontaneous acute ICH were included. Exclusion criteria were incidental ICH on head CT, non-stroke activation patients, or secondary causes of ICH.

### Outcomes

The primary outcomes were pre- to post-intervention differences in the time from CT scout image to administration of antihypertensive medication and anticoagulation reversal.

Secondary outcomes included pre- to post-intervention comparison of: door to treatment for administration of antihypertensive medication and anticoagulation reversal, CT scout to stroke orderset activation, hospital LOS, discharge disposition, and mortality. Exploratory subgroup analysis was also performed for patients who were cared for in the ED by the night telestroke or teleneurocritical care provider, as compared to the daytime in-person provider.

### Statistical Analysis

Statistical analyses were performed on all patients included in the study. Categorical and ordinal data were analyzed using χ2, Fisher’s exact test, or Mann-Whitney U test. Continuous data were analyzed by t-tests or Mann-Whitney U tests. All hypothesis tests, where applicable, were 2-sided. The author who performed statistical analyses was blinded to the hospital type, time of patient presentation, pre-versus post-intervention patient groups, and then unblinded for final comparisons.

Statistical software utilized were IBM SPSS Statistics Version 28 (IBM, Armonk, NY) and GraphPad Prism 7 (GraphPad Software, La Jolla, CA).

## RESULTS

### Patients

A total of 381 patients were assessed for eligibility, and 226 patients (108 pre-intervention and 118 post-intervention) met all inclusion criteria (Figure 2). Patient demographics were similar (Table 1). Systolic blood pressure (SBP) on admission between pre- and post-interventions groups were not different (median, IQR: 173, 158-208 mmHg vs. 176, 147-206 mmHg; p=0.82). The pre- and post-intervention median NIHSS scores were 10 (IQR 6-21) vs. 10 (IQR 5-18; p=0.25); ICH scores were 2.0 (IQR 0-3) vs. 1.0 (IQR 1-3; p=0.90), respectively (Table 1).

**Table 1.** Pre- and post-intervention of ICH feedback patient demographic and clinical characteristics.

|  | Pre-Intervention (n= 108) | Post-Intervention (n=118) |
| --- | --- | --- |
| Age (median, IQR) | 68, 56-77 | 69, 53-77 |
| Sex (% Female) | 57% | 53% |
| Race (%) |  |  |
| White | 86.1% | 80.5% |
| Black | 92.6% | 0.0% |
| Asian | 1.9% | 4.2% |
| Native American or<br>Hawaiian | 2.8% | 11.9% |
| Unable to determine | 8.3% | 3.4% |
| Daytime presentation (%) | 59.3% | 61.0% |
| Presented to non-CSC (%) | 63.0% | 57.6% |
| SBP at ED Presentation<br>(median mmHg, IQR) | 173, 158-208 | 176, 147-206 |
| DBP at ED Presentation<br>(median, IQR) | 102, 72-141 | 103, 76 - 134 |
| NIHSS (median, IQR) | 10, 6-21 | 10, 5-18 |
| ICH Score (median, IQR) | 2, 0-3 | 1, 1-3 |

Abbreviations: CSC, comprehensive stroke center; DBP, diastolic blood pressures; ED, emergency department; ICH, intracerebral hemorrhage; IQR, interquartile range; NIHSS, national institutes of stroke scale, SBP, systolic blood pressure.

**Figure 2.**
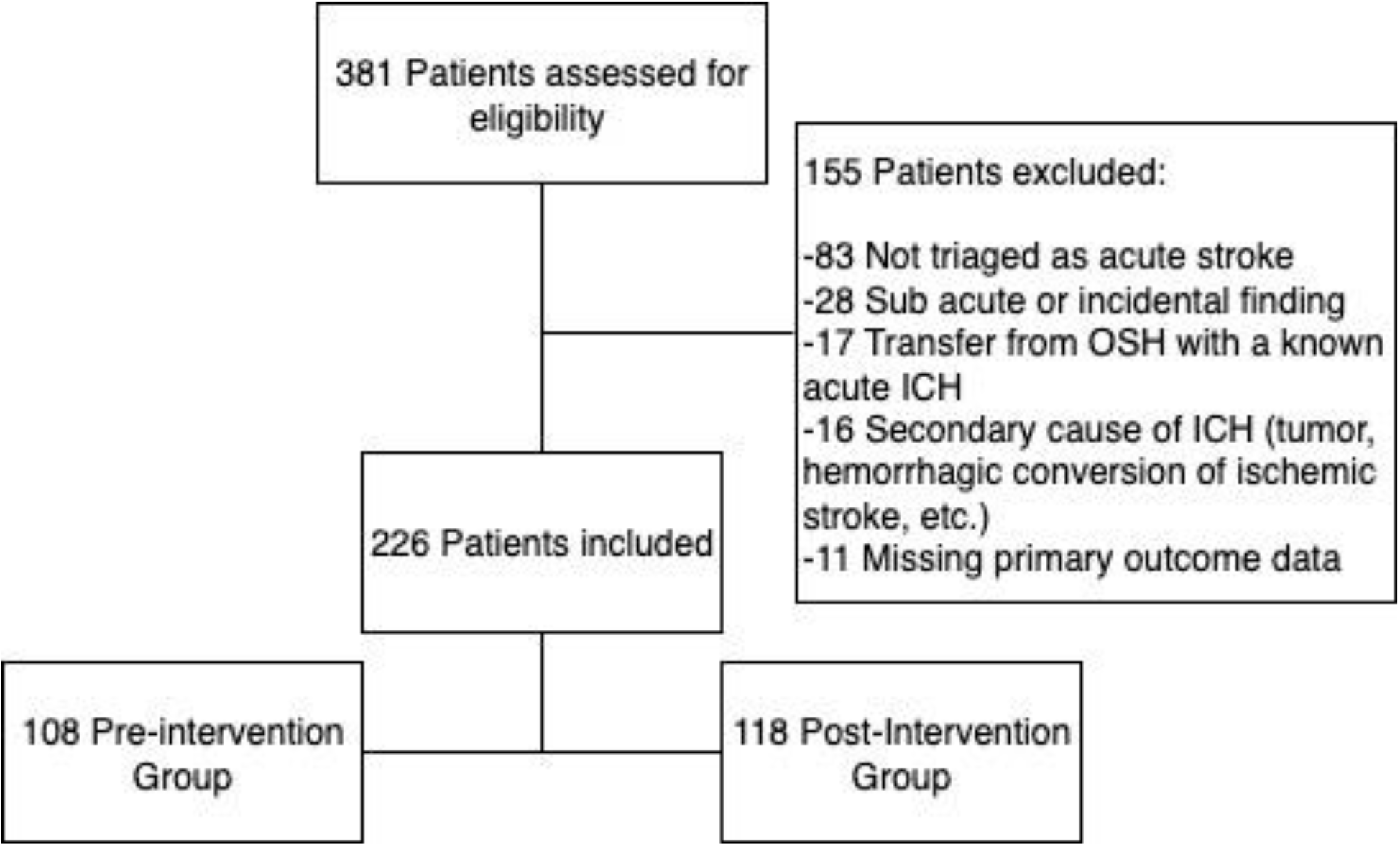
Patient eligibility and pre- and post-intervention groups flow diagram.

**Figure 3.**
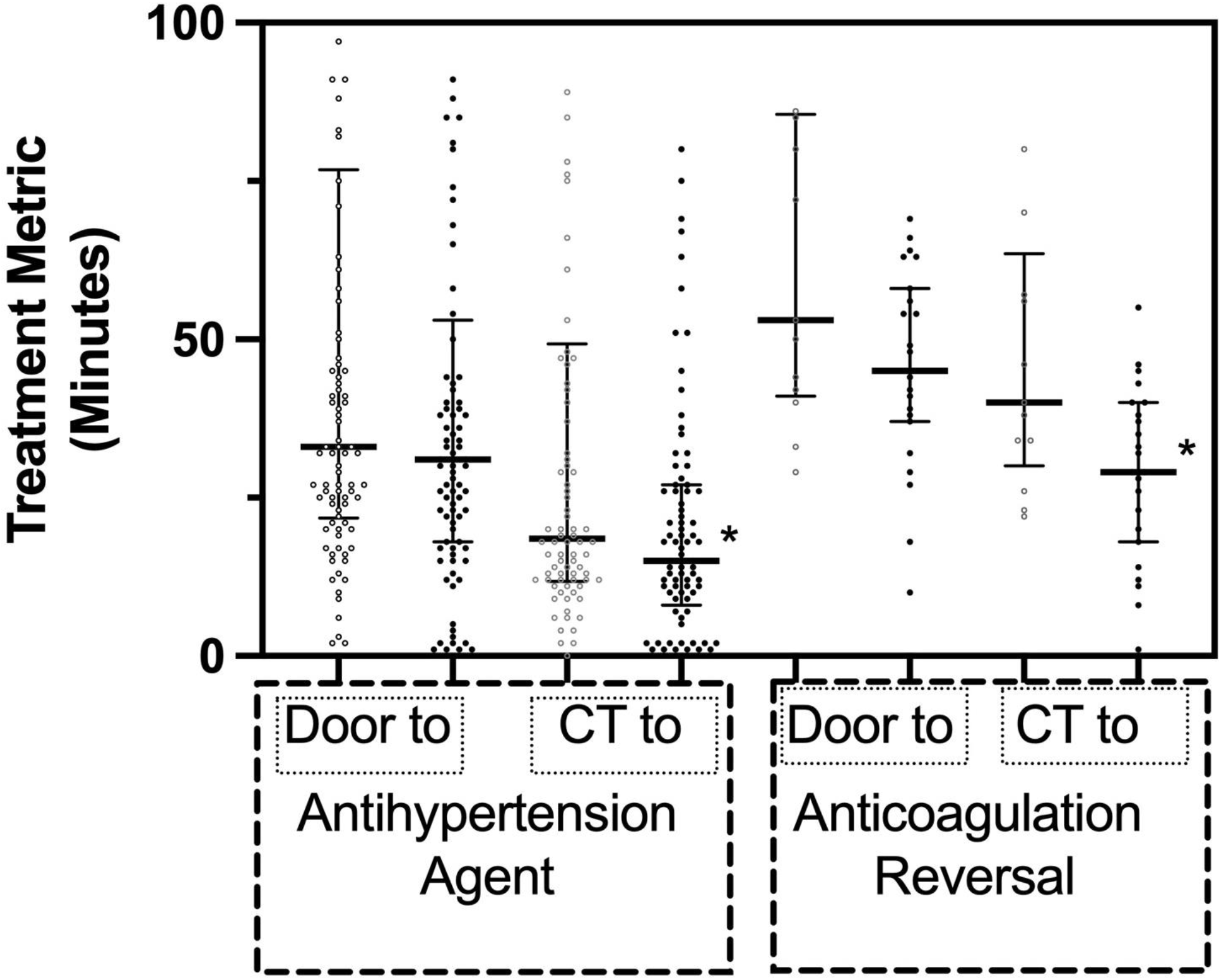
Treatment metrics for patients with spontaneous intracerebral hemorrhage (ICH) pre- and post-ICH feedback intervention. Door to treatment and CT scout image to treatment times are represented in pre-intervention (open, white circles) versus post-intervention (black squares). Thick horizontal line within each column represents the median and the thin shorter horizontal lines are the interquartile ranges. * p<0.05.

When an ICH was identified by the scout head CT, this served as time point zero for tailored ICH response protocols, specifically a bundled care ICH order set within the EMR. This orderset initiated specific ICH blood pressure goals, antihypertensive medications, anticoagulation reversal agents, and other treatment pathways, such as arterial line placement and neurosurgical consultation, in appropriate cases. In the post-intervention period, the use of the orderset increased from 54% to 96% (p=0.001). However, in the pre- to post-intervention groups, median door to orderset initiation was similar (23, IQR 16-31 minutes vs. 24, 18-42 minutes; p=0.48).

### Primary and secondary outcomes

The effect of the feedback intervention on the individual “door-to-treatment” metrics trended positively, yet its effect was only significant in “scout CT-to-treatment” metrics. Median door to antihypertensive treatment pre- to post-intervention was unchanged (43, IQR 33-77 min vs. 36, 21-58 min; p=0.78). Conversely, when the start of time to treatment was the CT scan, then the intervention was associated with faster median time to antihypertensive treatment (21, IQR 23-52 min vs. 14, 7-26 min; p=0.0012; Figure 2). Median door-to-anticoagulation reversal administration pre- to post-intervention was lower, but not statistically significant (53, IQR 41-86 min vs. 45, 37-58 min; p=0.07). When time zero was the scout CT scan, then the intervention did lead to significantly faster times (median, IQR: 40, 30-64 min vs. 29, 18-40 min, p=0.03).

Subgroup analysis was performed to evaluate the effect of the feedback intervention during the daytime in-person neurology coverage hours versus nighttime hours, which is covered by in-system teleneurology and teleneurocritical care services. During nighttime tele hours pre- to post-intervention there was reduction in median CT to antihypertensive administration time (32, IQR 20-84 min vs. 15, 11-29 min; p=0.012) and median anticoagulation reversal administration time (67, IQR 43-85 min vs. 45, 35-64 min; p=0.038, Figure 4). There was no difference pre- to post-intervention during daytime as well as nighttime hours door to antihypertensive treatment nor reversal agent administration (p>0.05). Similarly, there was no effect in daytime scout CT to antihypertensive treatment, reversal agent; p>0.05).

**Figure 4.**
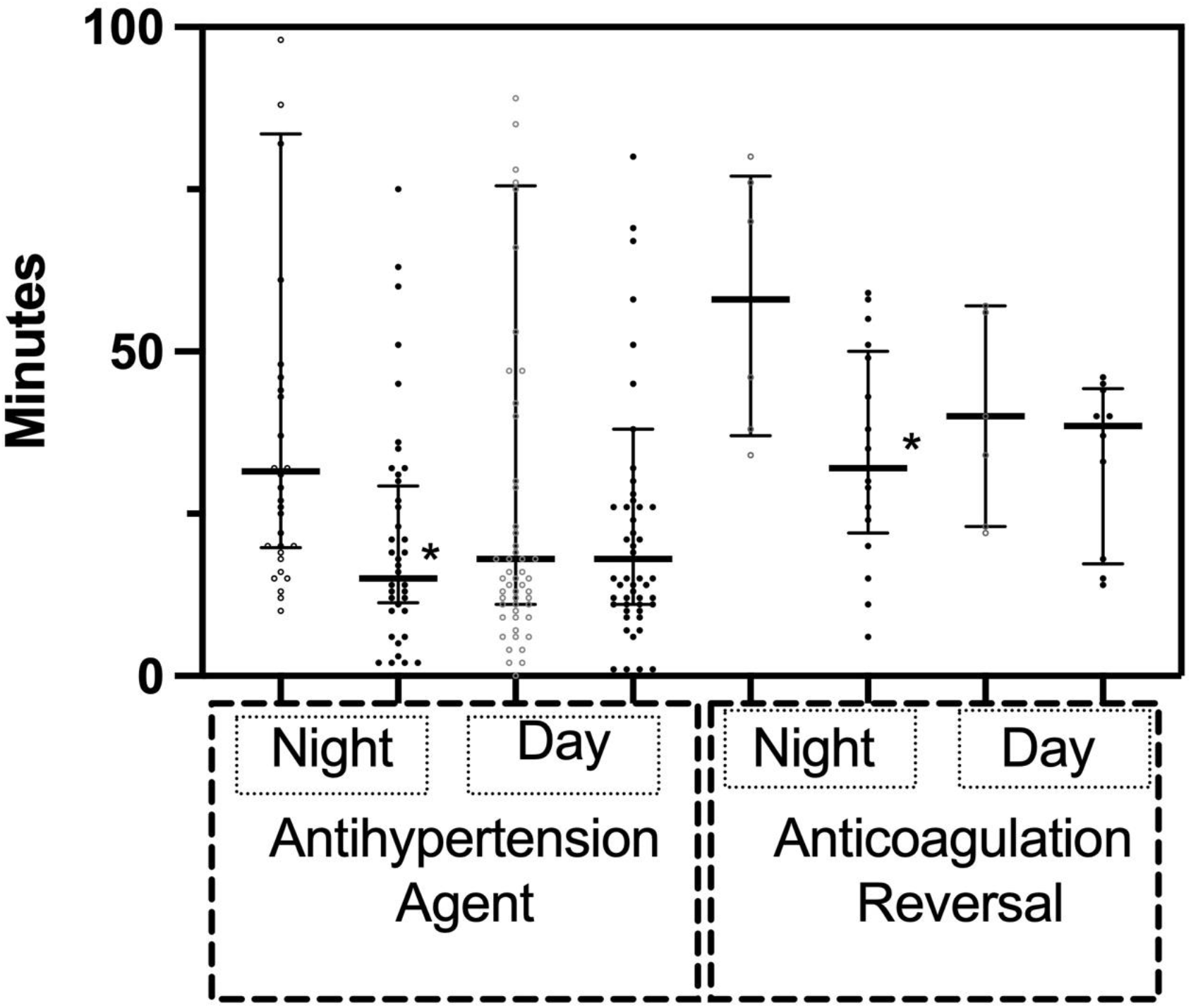
Treatment metrics for patients with spontaneous intracerebral hemorrhage (ICH) pre- and post-ICH feedback intervention subgrouped by nighttime versus daytime neurologist provider coverage. Metrics are represented as pre-feedback intervention (open, white circles) versus post-intervention (black squares). Thick horizontal line within each column represents the median and the thin shorter horizontal lines are the interquartile ranges. * p<0.05.

### Hospital utilization and outcomes

Hospital utilization metrics did not differ between groups, while disposition metrics did. Median hospital LOS was similar pre- and post-intervention (8, IQR 4-13 days vs. 7, 4-12 days; p=0.55). When patients were dichotomized to good discharge disposition, defined as acute rehab unit or home, versus poor (long term care facility, skilled nursing facility, or mortality) there was improvement in the percentage of patients who had a good outcome following the intervention (49.4 to 66.3%; p=0.025). When mortality was the binary outcome, there was no difference (pre n=3 patients, 3.5%, and post n=8 patients, 7.9%; p=0.23).

## DISCUSSION

Implementation of rapid feedback to provider teams caring for acute intracerebral hemorrhage treatment cases in the emergency department setting led to faster time to treatment. The effect was seen best by reducing CT head scout image to treatment times, rather than door to treatment, with an overall seven-minute reduction to antihypertensive treatment and eleven-minute reduction to anticoagulation reversal.

There are multiple reasons why faster time to treatment occurred primarily after the head CT imaging, rather than entry via the ED door. Door time to CT scanner processes for patients with acute stroke symptoms has already been optimized as a result of ischemic stroke workflows.(7)(8) Then, until the ICH becomes known by head CT images, the goal for blood pressure and magnitude for lowering cannot be realized, nor the pharmacist led preparation of anticoagulation reversal agents. Hence this is the time zone in which ICH specific interventions and process improvements are likely to yield effect.

The feedback intervention had a strong improvement effect on nighttime telestroke and teleneurocritical care acute management of ICH. Prior studies have revealed that nighttime treatment for stroke, particularly ischemic stroke, is slower and thus stands to improve more from targeted interventions.(11) Here, the nighttime-based median 17 and 22-minute reduction of time to antihypertensive drug administration and anticoagulation reversal, respectively, is clinically meaningful. The driver of such improvement could be enhanced team awareness of treatment metric goals at night, especially when pharmacy staffing is less dense, and a teleneurologist is covering multiple hospitals.

This study has limitations. Few patients had an indication for anticoagulation reversal agent administration, limiting statistical strength. Next, we excluded patients who did not receive a stroke code activation on presentation to the ED, since the time-sensitive protocols were not used in their care pathway. Third, the feedback intervention used here was rolled out over a 6-month period at different hospitals, so it is possible that movement of providers between hospital EDs during that period could bias treatment toward improvement even if the intervention had not yet been formally rolled out at the specific hospital’s ED. Finally, there may be interaction between increased familiarity of the ICH orderset as a result of its use and feedback, which in itself could have driven improvements.

Ultimately, directed process and feedback interventions to improve time sensitive treatment to ICH are needed and that used here may be useful for other hospital systems. Yet such feedback is resource intensive, and universal care protocols and treatment goals may not accommodate variations in hospital capabilities. Future studies may consider options for automated feedback, assessment of feedback effect for members working remotely via telehealth, and evaluation of long-term patient outcomes.

## CONCLUSION

A short interval ICH feedback intervention improves treatment times for antihypertensive medication administration and anticoagulation reversal following detection of ICH on CT head imaging. The standardized feedback template here may be useful for process improvements at other hospitals.

## Data Availability

Data is available with reasonable request.

